# Lung function trajectories in children with cystic fibrosis aged 3-17 years: impact of elexacaftor-tezacaftor-ivacaftor on lung function

**DOI:** 10.64898/2026.08.31.26361791

**Authors:** Brett P Dyer, Maddy Deery, Rebecca Heyman, Paul D Robinson, Claire E Wainwright, Peter D Sly, Robert S Ware, Tamara L Blake

## Abstract

**Background:** Elexacaftor-tezacaftor-ivacaftor (ETI) has been demonstrated to improve lung function in clinical trials; however, evidence describing effects on trajectories and whether long-term improvements are sustained (>1-year) is lacking. We estimated within-person lung clearance index (LCI) trajectories before and after ETI initiation, assessing changes in level and rate of change, alongside acute LCI change, up to three years after ETI initiation.

**Methods:** Prospective observational study of children at a tertiary hospital. Children aged 3–17 years with ≥2 LCI testing occasions (i) before and (ii) after starting ETI were used to describe lung function trajectories. Children with ≥1 pre-ETI and ≥1 post-ETI LCI occasion(s) were used to describe acute LCI change after ETI initiation. Age-adjusted LCI trajectories for time periods (i) before and (ii) after ETI initiation were estimated using linear mixed-effects models, and pre- and post-ETI LCIs were compared using paired Wilcoxon tests.

**Results:** Mean pre-ETI and post-ETI longitudinal changes in LCI were -0.007 (95% CI: -0.28, 0.27; n=35) and 0.12 (95% CI: -0.17, 0.41; n=20) turnovers per year, respectively. Before ETI initiation, 57% (30/53) of patients had an LCI≥7.1 turnovers (indicating impaired lung function), compared to 26% (14/53) post-ETI, with a median LCI difference of -0.70 (95% CI -0.84, -0.46; p<0.001) turnovers. Within-individual variability in LCI decreased post-ETI.

**Conclusions:** Our real-world data within a unique longitudinal study provide a comprehensive picture of ETI benefit by outlining not only acute improvement in LCI but maintained stability in LCI trajectories and improved LCI stability sustained up to three years post-initiation.

## Background

Cystic fibrosis transmembrane conductance regulator (CFTR) modulator treatment improves CFTR protein function and/or quantity at the epithelial surface, with well-recognised improvements in health outcomes [1-3]. More recently, a triple-combination CFTR modulator, elexacaftor–tezacaftor–ivacaftor (ETI), has become widely available, with benefits noted in a broad range of CFTR variants [4-7]. In Australia, ETI became reimbursed and available to pwCF aged 12 years and older in April 2022, children aged 6 to 11 years in May 2023, and children aged 2-5 years in August 2024.

Lung clearance index (LCI), as measured by multiple breath washout (MBW), is recognised as the gold standard for measuring early CF lung disease progression in clinical trials and the effects of interventions due to its much stronger feasibility across age ranges and ability to detect peripheral airway changes resulting in increased ventilation inhomogeneity, requiring far smaller sample sizes than FEV_1_ [8]. Longitudinal MBW testing has provided unique insights into the development and progression of CF lung disease by outlining progressive decline in lung function trajectories (or worsening of ventilation inhomogeneity) from preschool through to adolescent years [9, 10].

LCI is now routinely used as an efficacy endpoint in CFTR modulator clinical trials, with short-term population mean reductions in LCI between -1.71 and -2.29 turnovers observed post-ETI initiation in 6-to 11-year-olds [11-13]. Similar findings are being seen in clinical (or real-world) settings, with a Danish study of 131 children aged 6-17 years demonstrating a reduction/improvement in mean LCI of 1.7 (95% CI: 1.2 – 2.1) turnovers between pre-ETI and 12 months post-ETI LCI values [14]. However, evidence on within-person lung function trajectories and longer-term changes in LCI, greater than one-year after ETI initiation, remains limited.

The Early Life Origins of CF (ELO) study is a unique longitudinal observational study which commenced in 2020 and focused on longitudinal changes in lung function, including annual MBW testing from the age of 3 years. The present study describes average within-person lung function trajectories pre- and post-ETI initiation in a cohort of ELO children with CF aged 3 to 17 years, from the largest CF clinic in the Southern Hemisphere, located at a tertiary paediatric hospital in Brisbane, Australia. We aimed to describe lung function trajectories (as mean within-patient rate of change in LCI), measured using LCI, before and after ETI initiation, and to assess whether any short-term changes in LCI were sustained for up to 3 years. We also examined whether the changes in LCI after ETI initiation were heterogeneous according to age, sex, genotype, and pancreatic sufficiency.

## Methods

### Study design and participants

This prospective observational study involved children with verified CF (genotype confirmed), aged <18 years at the time of ETI initiation, who participated in the ELO study between January 2020 and March 2025. All children who attended CF clinics at the Queensland Children’s Hospital, Brisbane, Australia, were approached to participate in the study. Written informed consent was obtained from parents. The study was approved by the local ethics committee (HREC/19/QCHC/51727).

All available data were screened for inclusion in three datasets for analysis. The first cohort, referred to as the “pre-ETI” cohort, consisted of participants with at least two technically acceptable MBW LCI measurements before being prescribed ETI. This group was used to assess lung function trajectories before ETI therapy. The second cohort, referred to as the “change” cohort, consisted of participants with at least one pre-ETI and at least one post-ETI LCI measurement. This group was used to describe the acute change in lung function after ETI initiation and to investigate factors that may influence this. The third cohort, referred to as the “post-ETI” cohort, consisted of participants with at least two LCI measurements after being prescribed ETI. This group was used to assess lung function trajectories following ETI therapy. Children who were on ivacaftor before ETI were excluded, as ivacaftor (a component of ETI) is effective for their gating mutation and switching to ETI was not expected to lead to any further benefit in respiratory function.

### Variables and data collection

MBW was performed in all children aged ≥3 years at approximately yearly intervals during routine clinic visits (EXHALYZER® D, Eco Medics) as per consensus recommendations, reporting LCI as the mean of at least two technically acceptable trials [15, 16]. All test occasions were analysed in the latest version of software (Spiroware v3.3.1) to ensure correction for a recent cross-talk sensor error [17, 18]. An LCI greater than or equal to 7.1 turnovers was used to define abnormal lung function across the age ranges tested [19, 20]. Height, weight, pancreatic sufficiency status (defined as pancreatic insufficient based on low faecal elastase level), clinical status (including information on recent pulmonary exacerbations) and microbiology results were documented after review of electronic medical records. Persistent infection was defined as either three positive samples, representing ≥50% of samples, within the preceding 12-month period or chronic use of anti-bacterial therapy (e.g. anti-*Pseudomonas aeruginosa* antibiotics) [21, 22]. Age was defined as the age at the time of ETI initiation, and groups (3-5, 6-11, 12-17) were chosen based on ETI rollout, with ages 6-8 years and 9-11 years separated to further examine heterogeneity.

### Analysis

Linear mixed-effects regression was used to estimate the average rate of change in LCI per year (i) in the time period before patients started ETI and (ii) in the time period after patients started ETI. Random effects for the intercept and time (years) were used to account for dependence between repeated measures. As both LCI and timing of ETI initiation are strongly affected by age, models were adjusted for age on the day of the first LCI measurement. Unadjusted results are also presented. Another model, including age, pre-ETI modulator (lumacaftor/ivacaftor or no CFTR modulator) and the interaction between the pre-ETI modulator and time, was used to examine whether pre-ETI trajectories differed according to previous CFTR modulator among children of the same age.

Paired Wilcoxon tests were conducted to assess the marginal and within-group differences between the distribution of the last LCI measurements taken before ETI and the distribution of the first LCI measurements after the start of ETI. The proportion of patients with a clinically meaningful change in LCI of >10% was also calculated [23]. Kruskal-Wallis (>2-group comparison) and Mann-Whitney U tests (2-group comparison) were used to compare the differences in the change in LCI from before ETI initiation to after ETI initiation between subgroups. McNemar’s test was used to assess the difference in the proportion of participants with LCI ≥ 7.1 turnovers before and after ETI initiation. Tests were only conducted for subgroups containing five or more patients. Data were analysed using R version 4.4.0 [24].

## Results

### Study samples

As shown in Figure 1, 167 children from the ELO study, aged between 3 and 17 years, completed at least one technically acceptable MBW measurement between January 2020 and March 2025. Sixty children did not meet CFTR modulator criteria. Thirty-five, 53, and 20 patients, respectively, met eligibility criteria, based on the number of MBW test occasions before and after ETI initiation, for the “pre-ETI”, the “change”, and the “post-ETI” cohorts.

**Figure 1.**
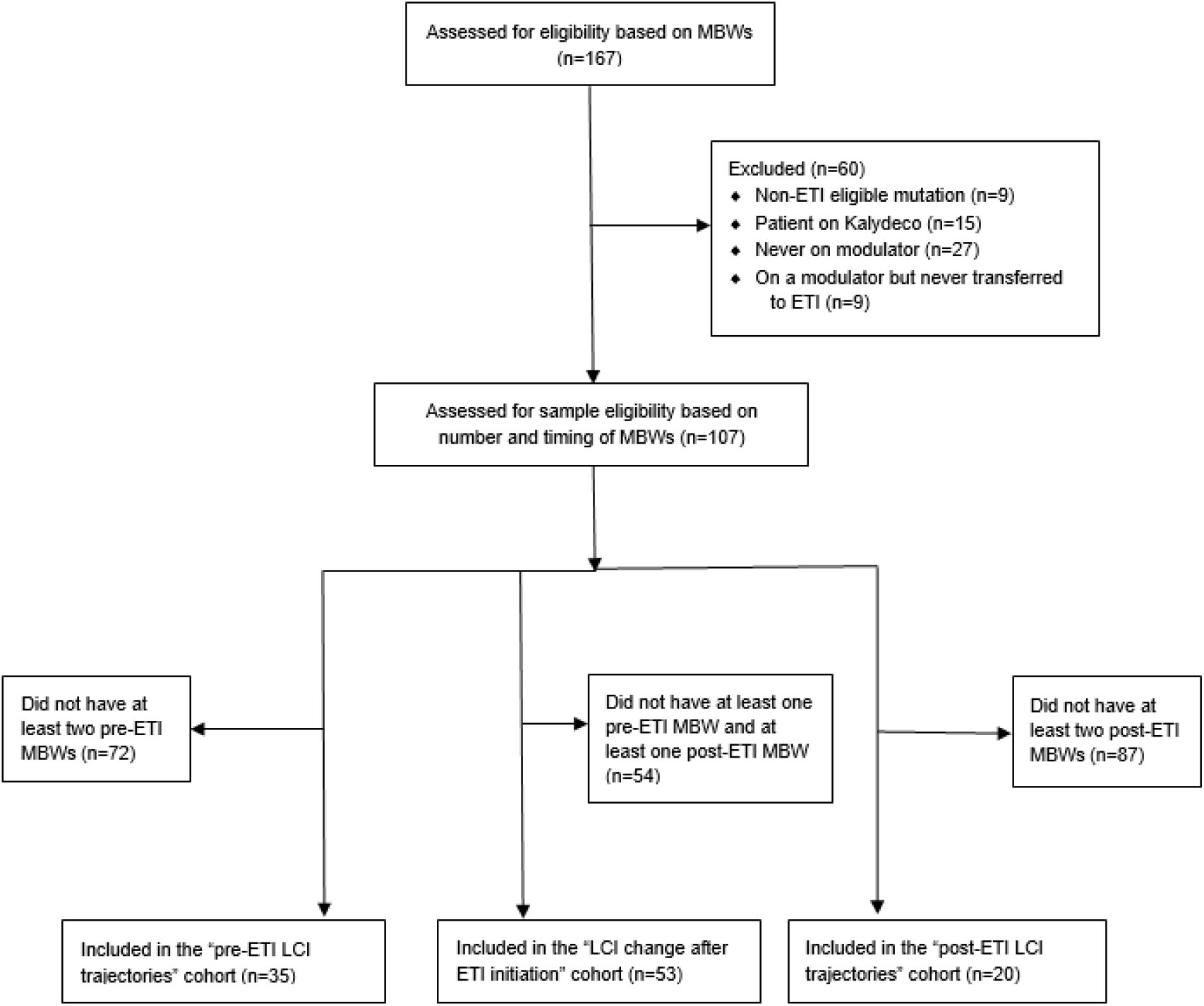
Participant flow diagram

### Baseline sample characteristics

The baseline characteristics of patients in each sample were similar (Table 1). As expected, the median age in the “pre-ETI” cohort (7.5 years) was lower than in the “change” cohort (9.3 years), which was lower than in the “post-ETI” cohort (10.8 years).

**Table 1.**
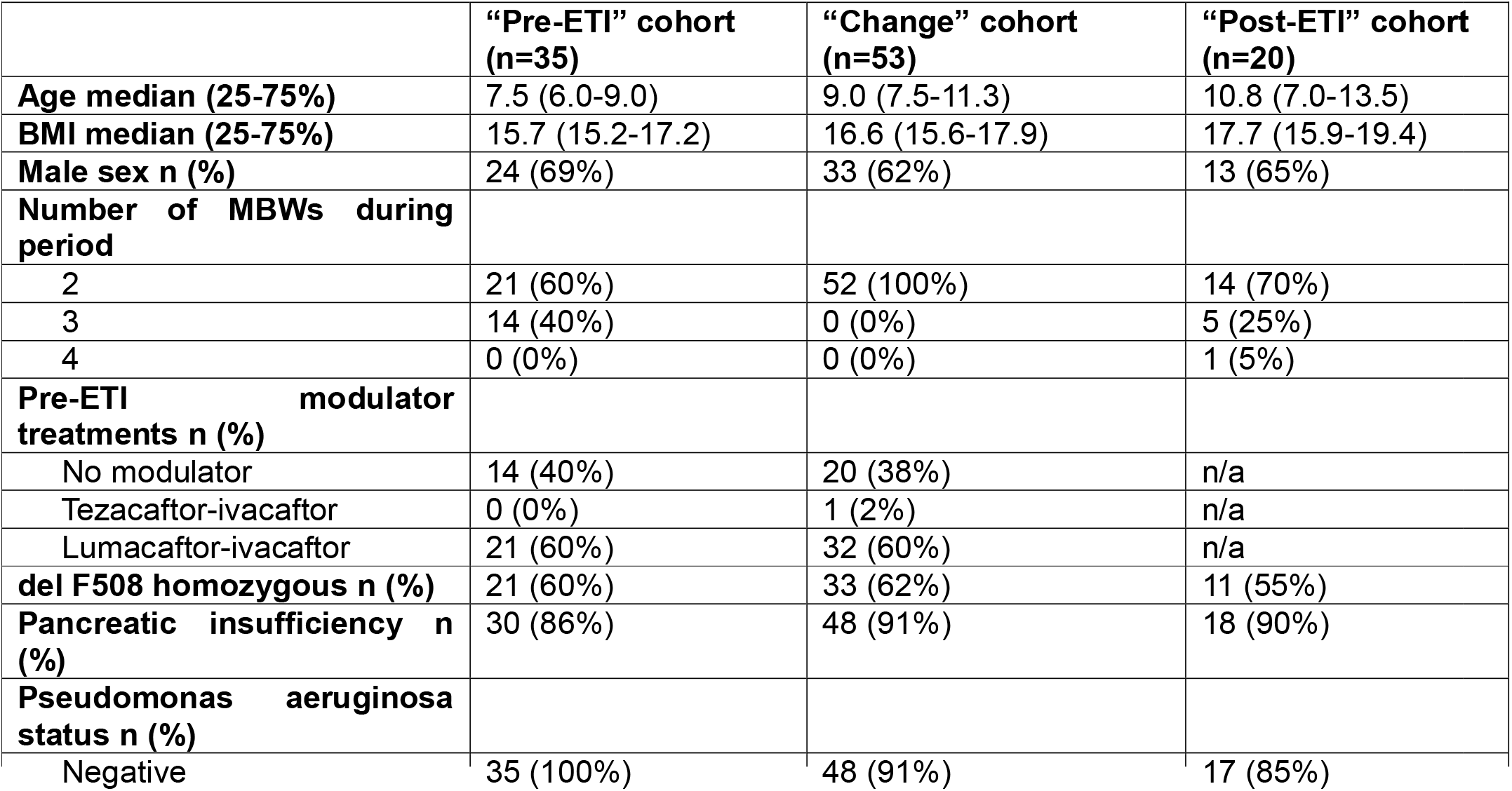
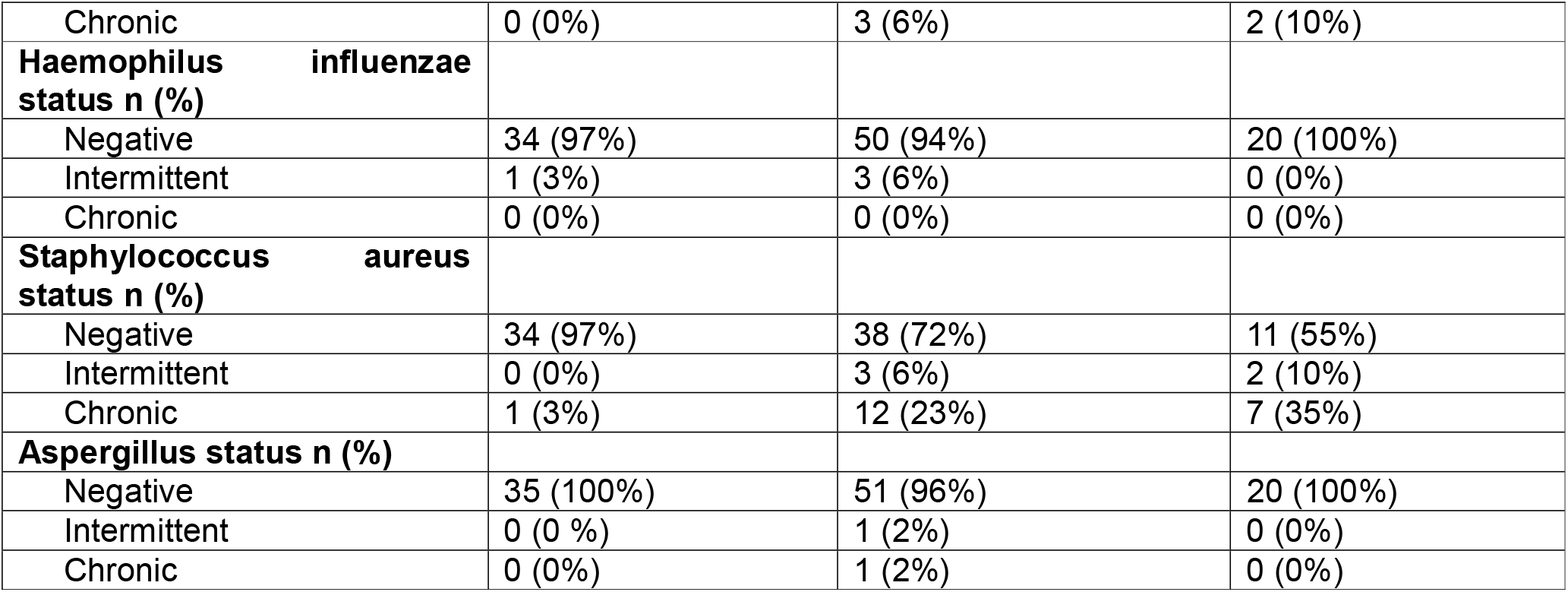
Baseline sample characteristics.

### Trajectories pre-ETI initiation

Individual trajectories of LCI pre-ETI initiation are shown in blue in Figure 2. In the age-adjusted analysis, the average change in LCI was -0.0066 (95% CI: -0.28, 0.27) turnovers·year^-1^. Thus, there was not evidence of a change in LCI among people of the same age during this period. The standard deviations of the intercept random effects, the slope random effects, and residuals were 1.31 turnovers, 0.63 turnovers·year^-1^, and 0.67 turnovers, respectively. This indicates substantial between-individual variability in LCI values at time zero and in LCI trajectories, as well as substantial within-individual variability over time. In the unadjusted analysis, the average increase in LCI was 0.012 (95% CI: -0.28, 0.30) turnovers·year^-1^, again not providing evidence of an increase or decrease in LCI during this time period. From the 84 MBWs performed in this time period, 41 (49%) measured an abnormal LCI.

**Figure 2.**
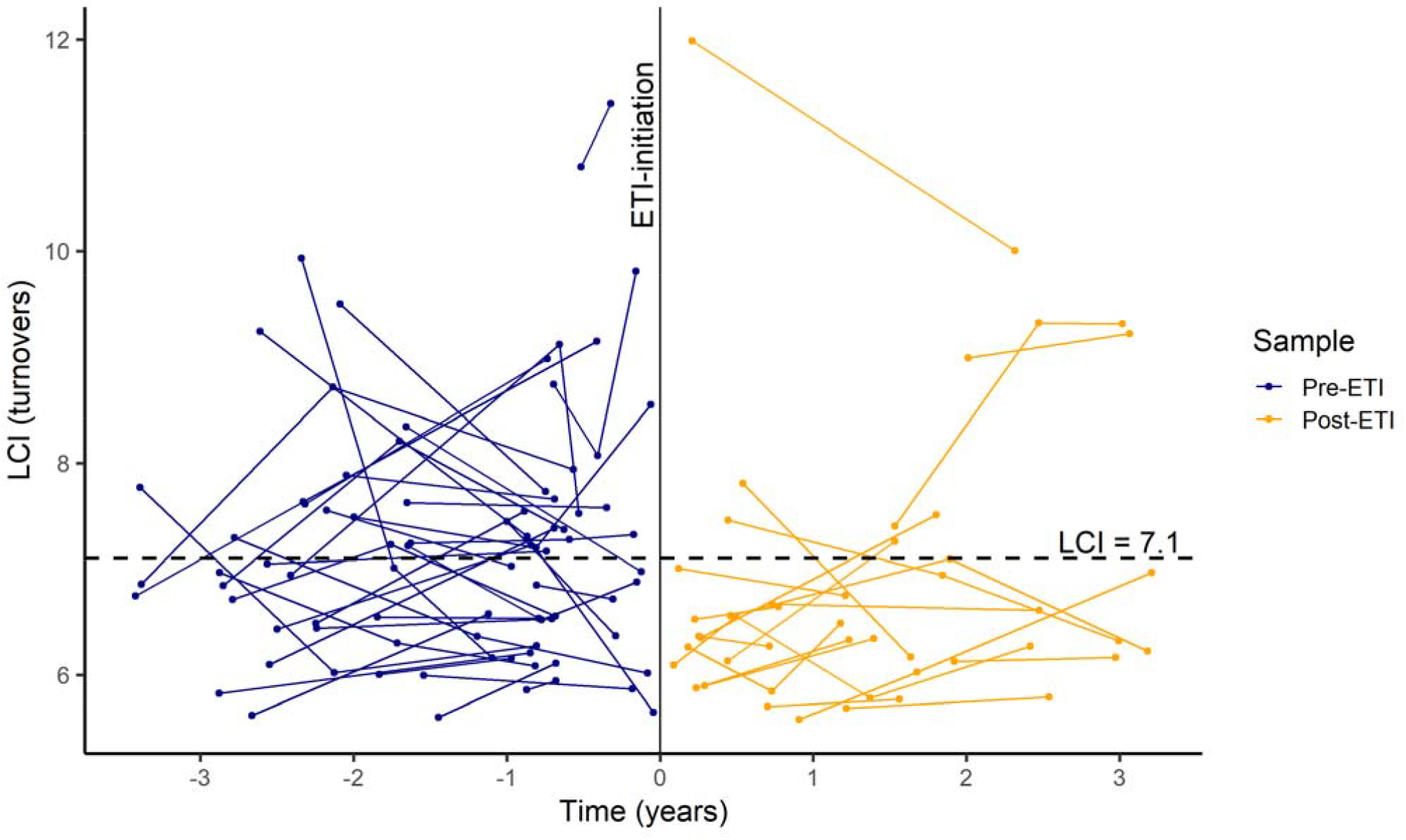
Lung function (LCI) trajectories (i) pre-ETI treatment initiation (coloured blue) and (ii) post-ETI initiation (coloured orange). Footnote: LCI abnormality is defined as 7.1, such that a value above 7.1 indicates abnormal lung function. Grey dashed lines are used to connect data points across time zero for the three patients who contributed to both the pre-ETI (objective 1) and post-ETI (objective 3) trajectory samples.

In the model including the interaction term between time and pre-ETI modulator, the average annual change in LCI was estimated to be 0.30 (95% CI: -0.15, 0.75). The difference in LCI at time zero between those who were on lumacaftor-ivacaftor, compared to those who were not previously on a CFTR modulator, was estimated to be -0.50 (95% CI: -1.57, 0.56), and the difference in the rate of change in LCI between those on lumacaftor-ivacaftor, compared to those who were not previously on a CFTR modulator, was estimated to be -0.46 (95% CI: -1.02, 0.11). Thus, there was not clear evidence of a difference in LCI at time zero or pre-ETI trajectory based on whether the patient was previously on lumacaftor-ivacaftor or if they were not on a CFTR modulator, with 95% CI within the normal between-session repeatability of the MBW in this setting [23].

### Change in lung function post-ETI initiation

Figure 3 shows a pattern of decreasing LCI from the last measurement taken pre-ETI initiation to the first measurement post-ETI initiation. Prior to ETI therapy, 30 of 53 (57%) patients had an abnormal LCI, compared to 14 of 53 (26%) post-ETI therapy (p<0.001). Further, 46 of 53 children (87%) had a decrease in LCI following commencement of ETI, of which 26 of 53 (49%) had a clinically significant change (i.e. >10% decrease). The median (IQR) LCI before treatment was 7.2 (1.4) turnovers compared to 6.5 (1.0) turnovers after treatment (median difference -0.70 turnovers, 95% CI -0.84, -0.46; p <0.001). Thus, there is strong evidence that, on average, LCI values were lower after starting ETI therapy, with 31% more patients having LCI values within the normal range once initiated on ETI therapy.

**Figure 3.**
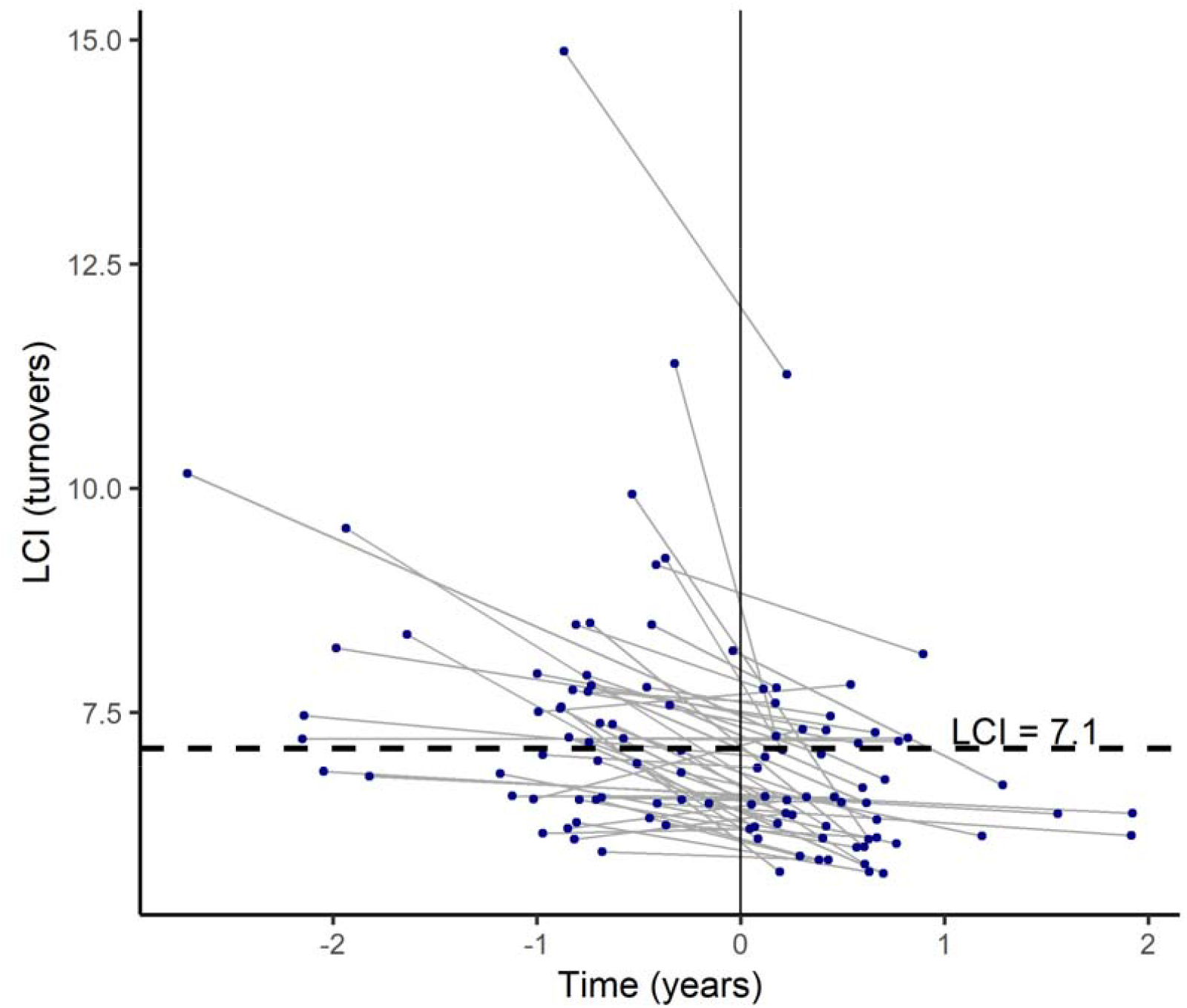
The change in lung function (LCI) between the last MBW pre-ETI therapy initiation and the first MBW post-ETI therapy initiation. Footnote: Upper limit of normal for LCI is defined as 7.1 turnovers (vertical line), and values of LCI above this threshold are considered abnormal.

### Trajectories post-ETI initiation

Individual trajectories of LCI post-ETI initiation are shown in orange in Figure 2. In the age-adjusted analysis, the average increase in LCI was 0.12 (95% CI: -0.17, 0.41) turnovers·year^-1^. Thus, there was not clear evidence of a change in LCI among people of the same age during this period. The standard deviations of the intercept random effects, the slope random effects, and residuals were 1.57 turnovers, 0.54 turnovers·year-1, and 0.36 turnovers, respectively. Again, this indicated substantial between-individual variability in LCI values at time zero and in LCI trajectories. The within-individual LCI standard deviation was lower in this post-ETI model than in the model describing pre-ETI trajectories (0.36 vs 0.67, respectively), which could suggest greater LCI stability following ETI initiation. The median follow-up time was 2.0 years (25-75%: 1.4 – 3.0). In the unadjusted analysis, the average increase in LCI was 0.12 (95% CI: -0.18, 0.42) turnovers·year^-1^, again not providing evidence of a clear increase or decrease in LCI during the time period following ETI initiation. From the 47 MBWs performed in this time period, 11 (23%) had an abnormal LCI.

### Factors associated with lung function pre- and post-ETI initiation

Table 2 summarises the median LCI from the last MBW before starting ETI and the first MBW after starting ETI, grouped according to sex, age group when starting ETI, delF508 genotype, and pancreatic sufficiency. Median LCI decreased in all subgroups post-ETI initiation. All subgroups showed statistically significant LCI improvement (all p<0.05), except pancreatic-sufficient individuals (n=5; p=0.13), likely reflecting low power.

**Table 2.** A summary of LCI measurements from the last MBW before starting ETI and the first MBW after taking ETI, grouped according to patient characteristics.

| Subgroups | Median pre-ETI LCI (IQR) | Median post-ETI LCI (IQR) | Median change in LCI (IQR) | p (pre vs. post) | p (between subgroups)<sup>†</sup> |
| --- | --- | --- | --- | --- | --- |
| **Age (yrs) when starting ETI** |  |  |  |  |  |
| 3 to 5 (n=3) | 9.2‡ | 7.2‡ | 2.1 | * | 0.36 |
| 6 to 8 (n=24) | 7.2 (6.8-7.8) | 6.4 (6.1-7.1) | 0.66 (0.28-1.02) | <0.001 |  |
| 9 to 11 (n=18) | 7.0 (6.5-7.9) | 6.5 (6.1-6.9) | 0.64 (0.08-1.04) | 0.0024 |  |
| 12 to 17 (n=8) | 7.0 (6.8-7.6) | 6.4 (6.1-6.7) | 0.59 (0.32-0.87) | 0.023 |  |
| **Sex** |  |  |  |  |  |
| Male (n=33) | 7.2 (6.6-8.4) | 6.5 (6.1-7.2) | 0.71 (0.15-1.27) | <0.001 | 0.39 |
| Female (n=20) | 7.3 (6.7-7.7) | 6.4 (6.1-7.1) | 0.53 (0.30-0.93) | <0.001 |  |
| **Previous CFTR modulator** |  |  |  |  |  |
| No CFTR modulator (n=20) | 7.1 (6.5-8.0) | 6.4 (6.1-7.2) | 0.58 (0.27-1.12) | <0.001 | 0.68 |
| Lumacaftor-ivacaftor (n=32) | 7.4 (6.7-8.1) | 6.5 (6.2-7.1) | 0.71 (0.30-1.03) | <0.001 |  |
| **delF508 genotype** |  |  |  |  |  |
| Homozygous (n=33) | 7.5 (6.8-7.9) | 6.5 (6.2-7.1) | 0.71 (0.15-1.07) | <0.001 | 0.74 |
| Heterozygous (n=20) | 7.1 (6.5-8.0) | 6.4 (6.1-7.2) | 0.58 (0.27-1.12) | <0.001 |  |
| **Pancreatic sufficiency** |  |  |  |  |  |
| Sufficient (n=5) | 7.9 (6.8-8.5) | 6.4 (6.1-6.5) | 0.63 (0.48-2.49) | 0.13 | 0.7 |
| Insufficient (n=48) | 7.2 (6.5-7.8) | 6.5 (6.1-7.2) | 0.70 (0.15-1.07) | <0.001 |  |

## Discussion

To our knowledge, this is the first study to describe changes in patient-level trajectories of LCI and LCI variability pre- and post-initiation of ETI, alongside the acute LCI changes observed. Our results extend current understanding in the literature by outlining beneficial effects on LCI variability and ongoing stability in LCI trajectories out to three years post-ETI initiation, while confirming the real-world beneficial short-term impact of ETI on LCI.

We provide updated LCI trajectory data outlining an improvement in LCI trajectories, comparing our data with other published cohorts over time. Historically, registry data have always shown lung function worsens with increasing age, despite progressive improvement in general health outcomes and survival for pwCF, with abnormal spirometry persisting into adults [25, 26]. Mitigating this deterioration has been a long-standing goal of the CF community, and the hope had been that earlier access to CFTR therapies would achieve this over time. Initial LCI trajectories published in the literature, by Frauchiger *et al*., showed worsening LCI trajectories during the paediatric age range, beginning in school-aged children with more pronounced deterioration in older ages: an average increase in LCI of 0.21 (95% CI: 0.07–0.35) turnovers·year^-1^ in 6-11-year-olds, and 0.41 (95% CI: 0.27–0.54) turnovers·year^-1^ in 12–18-year-olds [10]. Our finding that the average pre-ETI LCI trajectory in the present study of -0.0066 (95% CI: -0.28, 0.27) turnovers·year^-1^ in 6–17-year-olds therefore represents a significant addition to the literature as the first evidence of achieving that goal from the preschool age range. Earlier evidence that this might be achievable was suggested in the LCI trajectories described by Stanojevic et al. in a North American cohort, where LCI trajectories flattened during later school-age years [27]. In the Stanojevic et al cohort, the proportion of subjects prescribed mucus clearance therapies and CFTR modulators increased between the preschool and school-age years as trajectories flattened [27]. Lumacaftor-ivacaftor, the most common CFTR modulator that our cohort was prescribed at baseline (prescribed in 62% of our cohort), has also been shown to improve MBW trajectories in children [28].

We also describe the novel finding that ETI improved stability in lung function, as assessed by MBW, over time. Within-individual variability of LCI values decreased in our dataset comparing pre-ETI and post-ETI periods.

By anchoring our data to the date at which ETI commenced, rather than age range as used in previous cohorts, we are able to more clearly show the effects of ETI on these important measures of clinical response within the same analysis: LCI trajectory over time, within-individual standard deviation of LCI values, and acute change in LCI value. We add to the existing evidence outlining acute LCI improvement following ETI initiation in children with CF aged 3 to 17, consistent with evidence from clinical trials[12, 29-31] and two other recent real-world studies [14, 32]. In 131 Danish children aged 6 to 17, mean LCI decreased from 9.05 turnovers (95% CI: 8.04, 10.07) pre-ETI to 7.4 (95% CI: 6.37, 8.43) 12 months after starting ETI [14]. As in our cohort, the majority (≈80%) of the Danish cohort were on an existing CFTR modulator, and their larger mean LCI improvement likely reflects their higher baseline LCI values [14]. In a largely CFTR modulator-naïve (≈75%) cohort of 59 Swiss children aged 4 to 18, median LCI decreased from 7.9 turnovers (IQR: 2.7) to 6.4 (IQR: 0.9) 12 months after starting ETI [32]. The magnitude of LCI improvement was more comparable to our values (median LCI before ETI was 7.2 turnovers (IQR: 1.4) vs. 6.5 (IQR: 1.0) after treatment).

Finally, we reinforce the long-term effectiveness data outlined in clinical trials in our real-world data. Sustained improvements in lung function following ETI initiation have been demonstrated up to 24 weeks in children aged 2 to 5 years [31] and up to 192 weeks in children aged 6 to 11 years [29, 30] in phase 3 clinical trials. We demonstrated both a short-term improvement in LCI following ETI initiation and an ongoing ability to maintain a stable LCI trajectory over time, sustained over follow-up of up to three years.

This study has key strengths. Firstly, whilst short-term changes in LCI have been described in observational studies, alongside population-average improvements over the first 12 months, this is the first observational study (to our knowledge) to describe LCI trajectories for up to three years post-ETI initiation. Secondly, clinical trials historically have lacked pre-ETI trajectory data, typically only reporting from a single pre-ETI measurement. We know that LCI trajectory during early life is a significant predictor of abnormal LCI and spirometry in adolescence and adulthood [8, 10]. Together, this information provides unique insight into the extent to which lung function improvement may be sustained after starting ETI in real-world clinical settings. A further strength of this study is the size of the cohort. Queensland Children’s Hospital has a large State-based CF service (≈470 patients) spread across a very wide geography. Cohort size enabled both pre- and post-ETI average patient-level trajectories to be described, as well as LCI differences after starting ETI. Considerable MBW quality control expertise exists within the study team due to its role as a regional Central Over-Reading Centre (CORC) for international CF clinical trials employing MBW as an outcome measure.

This study has several limitations. First, data were collected at annual routine clinic visits rather than pre-specified timepoints; the median pre-ETI follow-up of 2.2 years may have been insufficient to detect a modest longitudinal LCI trend had one existed, and the wide confidence interval around the pre-ETI average annual change (-0.28 to 0.27 turnovers·year^−1^) reflects the substantial between-and within-individual MBW variability in this cohort. The pre-ETI trajectory analysis also does not control for the impact of other therapies used concurrently by the treating physician. The modest sample size and imbalance across some subgroups reduced the power to detect differences in the magnitude of change in lung function observed following ETI initiation between subgroups and our ability to demonstrate that the pre-ETI rate of change in LCI differed by pre-ETI CFTR modulator status. Larger studies or individual participant data meta-analysis can be used to examine whether there are predictors of treatment effect. Finally, these findings may have limited generalisability to younger children. Following the extension of ETI prescription in Australia in August 2024 to children aged 2-5-year-olds, only three children aged 3-5 (and no two-year-olds) provided data to allow for the comparison of LCI from before to after starting ETI. As more data becomes available, it will be important to continue the assessment of the safety and effectiveness of ETI in this age group.

In summary, the present study provides the first evidence that stabilisation of LCI trajectories is achievable over time for children aged 3 to 17, and that ETI initiation not only leads to an acute improvement in LCI (and the proportion with abnormality) but also to improved stability in lung function and ongoing ability to maintain stable trajectories of change up to 3 years post initiation. Of note, 26% of children remained above the LCI threshold of 7.1 turnovers following ETI initiation, identifying a residual higher-risk group for whom ongoing monitoring with MBW is warranted and for whom future studies should seek to identify predictors of incomplete treatment response.

## Supporting information

COI

## Data Availability

All data produced in the present study are available upon reasonable request to the authors

