## Supplementary material for "Lung function trajectories in children with cystic fibrosis aged 3-17 years: impact of elexacaftor-tezacaftor-ivacaftor on lung function": COI

| ICMJE DISCLOSURE FORM | |
| --- | --- |
| **Date:** | 2/4/2026 |
| **Your Name:** | Professor Claire Wainwright |
| **Manuscript Title:** | Lung function trajectories in children with cystic fibrosis aged 3-17 years: impact of elexacaftor-tezacaftor-ivacaftor on lung function |
| **Manuscript Number (if known):** | Click or tap here to enter text. |
| In the interest of transparency, we ask you to disclose all relationships/activities/interests listed below that are related to the content of your manuscript. “Related” means any relation with for-profit or not-for-profit third parties whose interests may be affected by the content of the manuscript. Disclosure represents a commitment to transparency and does not necessarily indicate a bias. If you are in doubt about whether to list a relationship/activity/interest, it is preferable that you do so.  The author’s relationships/activities/interests should be defined broadly. For example, if your manuscript pertains to the epidemiology of hypertension, you should declare all relationships with manufacturers of antihypertensive medication, even if that medication is not mentioned in the manuscript.  In item #1 below, report all support for the work reported in this manuscript without time limit. For all other items, the time frame for disclosure is the past 36 months. | |

|  | | | **Name all entities with whom you have this relationship or indicate none (add rows as needed)** | **Specifications/Comments (e.g., if payments were made to you or to your institution)** |
| --- | --- | --- | --- | --- |
| **Time frame: Since the initial planning of the work** | | | | |
| **1** | All support for the present manuscript (e.g., funding, provision of study materials, medical writing, article processing charges, etc.)  **No time limit for this item.** | | ☒ \| **None** \| Click the tab key to add additional rows. | |
| **Time frame: past 36 months** | | | | |
| **2** | | Grants or contracts from any entity (if not indicated in item #1 above). | ☐ \| **None** Yes – as listed in 5 and 9 \| Income on a per patient basis to institution derived from Pharmaceutical Studies conducted including GSK and Vertex pharmaceuticals | |
| **3** | | Royalties or licenses | ☒ \| **None** | |
| **4** | | Consulting fees | ☐ \| **None** Yes – as listed in 5 and 9 \| All honoraria paid to institution not for personal use. | |
| **5** | | Payment or honoraria for lectures, presentations, speakers bureaus, manuscript writing or educational events | ☐ \| **None** Yes \| All honoraria paid to institute, not for personal use. Vertex Pharmaceuticals: Sep2021 – Scientific Symposium – ERS International Congress 2021 – SHIFTing the boundary in rare diseases: spotlight on Cystic Fibrosis. Nov2021 – SHIFT Steering Committee – Medical Education Symposium Event Nov2021 – SHIFT Symposium. Aug2022 – SHIFT Symposium Steering Committee – medical education symposium. Aug2022 – CF Round Table – Practical considerations in managing patients with CF. Nov2022 – SHIFT Symposium Dec2022 – SHIFTing Focus Newsletter – Steering Committee. Mar2023 – Round Table – Managing Children with Cystic Fibrosis – Trikafta 6-11YO May2023 – Optimising the use of Trikafta in CF patients >6YO Nov2023 – SHIFT Symposium Dec2023 – HTA Scientific Webinar – Assessing the value of medicines: an introduction for clinicians. Mar2024 – TSANZ Conference – Conversation with CF experts on treating pre-school children with CFTR modulators: What have we learnt so far? | |
| **6** | | Payment for expert testimony | ☒ \| **None** | |
| **7** | | Support for attending meetings and/or travel | ☒ \| **None** | |
| **8** | | Patents planned, issued or pending | ☒ \| **None** | |
| **9** | | Participation on a Data Safety Monitoring Board or Advisory Board | ☐ \| **None** Yes – as listed \| All honoraria paid to institution not for personal use. Vertex Pharmaceuticals: Jun2021: PAED Medical Advisory Board Nov2021: Trikafta/Kaftrio Evidence Generation Global Advisory Board Nov2021-Oct2023 – Lead PI Services in connection with VX20-445-116 and VX20-445-119 Aug2022: CF <2YO Study Design Advisory Board Sep2022: Trikafta 6-11years old and the rare CFTR gene mutations indication – Advisory Board Feb2023: Ridgeline Study Investigator Meeting in Feb 2023 Feb2023-Feb2026: Lead PI Services in connection with VX22-445-122 | |
| **10** | | Leadership or fiduciary role in other board, society, committee or advocacy group, paid or unpaid | ☐ \| **None** Yes \| Deputy Editor Thorax 2020- Dec 2022, Associate Editor ongoing; \| Associate Editor Respirology; \| International Advisory Board Vertex Pharmaceuticals | |
| **11** | | Stock or stock options | ☒ \| **None** | |
| **12** | | Receipt of equipment, materials, drugs, medical writing, gifts or other services | ☒ \| **None** | |
| **13** | | Other financial or non-financial interests | ☒ \| **None** | |
| **Please place an “X” next to the following statement to indicate your agreement:** | | | | |
| ☒ | | I certify that I have answered every question and have not altered the wording of any of the questions on this form. | | |
